# Monitoring Military Pilots with Textile Sensors: Physiological Responses and Signal Quality Under Extreme Conditions

**DOI:** 10.1101/2025.09.03.25332904

**Authors:** René M. Rossi, Frederik Bauer, Andreas Stier, Dennis Bron, Simon Annaheim

**Affiliations:** Empa, Swiss Laboratories for Materials Science and Technology, Laboratory for Biomimetic membranes and textiles, St. Gallen, Switzerland; ETH, Department of Health Sciences and Technology, Zurich, Switherland; Armasuisse, Bern, Switzerland; Swiss Air Force, Aeromedical Center, Dubendorf, Switzerland

## Abstract

Military pilots are exposed to severe physiological challenges during their missions that may affect cognitive and physical performance. Therefore, continuous monitoring potentially provides critical insights about the impact of extreme condition exposure on the fitness for duty of military pilots. We applied a textile-based monitoring system during training sessions with pilot aspirants to investigate the impact of hypobaric hypoxia and high G-force exposures on the signal quality obtained for a 1-lead electrocardiogram (ECG) and chest excursions. The physiological variables considered were heart rate, heart rate variability, as well as respiratory frequency and respiratory amplitude. In general, 92% and 82% of the recorded ECG time segments during hypoxia and G-force exposure, respectively, were classified as plausible for further analysis. For respiratory data, 72% and 76% were classified as accurate for further data analysis and interpretation. Detailed information about signal quality was found to be critical for the assessment of physiological variables recorded in extreme conditions. Furthermore, the combination of various physiological signals allows for a more holistic interpretation of body responses, for the assessment of body tolerances and an early detection of possible physical and cognitive impairments.

## 1. Introduction

Military pilots are exposed to severe physiological challenges that may infringe on their physical and cognitive performance (1). Exposures such as high multiaxial g-forces and rotation, hypoxic conditions, radiation sources and high temperatures, or vibration and noise can lead to a reduction in their physical and mental performance, as demonstrated for severe hypoxic exposures (2).

This extraordinary physical strain requires increased physical and mental activities of the pilot to cope with stressors and to perform the flight tasks at a high level of accuracy. A higher activation level of pilots results in increased respiratory activity and an increase in heart rate (3), and a decrease in heart rate variability (HRV) (4). Furthermore, blood lactate concentration during hyperpnoea was found to be higher in hypoxia (5). Bustamante-Sánchez et al. (6) analyzed the psychophysiological responses of the participants in incremental hypoxia training (3 minutes at sea level, 8 minutes at 5,000 meters, and a maximum of 10 minutes at 7500 meters above sea level. They reported statistically significant decreases in blood oxygen saturation and increases in heart rate mean values during hypoxia. Also, a significant increase in the rating of perceived exertion was reported during hypoxia. Impairments of cognitive performance have also frequently been reported under high-altitude exposures (7).

Gravity-induced loss of consciousness (G-LOC) is caused by G acceleration stress and is one of the most life-threatening problems for pilots (8). A study from the UK Royal Air Force in 2017 revealed that 120 (14.8%) aircrew reported at least one episode of G-LOC, and in 260 cases (32.2%), at least one episode of almost loss of consciousness (A-LOC) occurred (9). The authors showed that the prevalence of reported G-LOC had decreased compared to an earlier survey in 2005 and concluded that this result might be due to the introduction of centrifuge training. The tolerance of the human body to acceleration forces depends on their magnitude, direction and duration, as well as individual factors like age or body weight (1).

To prevent such losses of consciousness, early warning algorithms based on electromyogram (EMG) measurements have been proposed (10). However, analyses using EMG sensors were restricted to training situations only. Other warning signs proposed are, for example, non-contact measurement systems detecting changes in eye blink rate and head position (11). A study employing multi-sensory data collection during flight simulations has established a more nuanced understanding of how pilots respond to sudden, critical exposures. The study analyzed subjective feedback from pilots and objective physiological data, creating a reliable index for distress levels and performance capabilities during critical flight phases (12,13). This method substantiates the need for a multidisciplinary approach to pilot evaluation, blending human factors with technology to enhance flight safety. However, the integration of well-validated bio-monitoring systems capable of recording changes in vital signs like heart rate, body temperature, respiration and body motion has not been considered so far (14). The measurement of physiological parameters, e.g. for the assessment of cognitive task load, has been proposed in several studies since the 1960’s, but the identification of reliable predictors for a reduction in mental or physical performance has been limited by the use of conventional physiological monitoring technologies (15). Recent studies investigated the possibility of using wearable sensors to assess vital signs like heart rate, respiratory rate or oxygen saturation (16). Such devices have been commonly used for several years in sports, but not for medical diagnostics due to higher demands on the accuracy and reliability of the systems. The use of multimodal data to predict soldiers’ health and readiness was repeatedly proposed by (17) while highlighting the need for an accurate assessment based on high-quality data.

Challenges of using wearable sensors to assess vital signs in aviation include the accuracy of the sensors used, their robustness to reliably record the vital signs in extreme conditions, as well as the integration in pilot suits and the interference with the pilot’s equipment (Shaw and Harrell 2023). Therefore, a continuous evaluation of the signal quality obtained from sensor recordings is crucial for the interpretation and assessment of the pilot’s readiness. Unfortunately, signal quality assessments are not adequately considered in recent literature (or at least not reported) (18,19). Data is missing about the impact of (severe) environmental working conditions and how this affects the data output of the systems and the accuracy of the further assessment. For field applications, the reliability and robustness of the system are critical aspects for the acceptance of new wearable technologies (17), as their ease of use and predicted usefulness (20). The relevance of the monitoring of vital signs and the validity of the predictive models used as early warning parameters has yet to be shown.

Among the different types of wearable health monitoring systems, textile-based sensors have emerged as an interesting technology due to their flexibility, softness, breathability, compatibility with pilot clothing, and conformation to each body location, which increases users’ comfort and acceptance, allowing continuous monitoring for extended durations. (21). However, as these systems are usually not directly attached to the skin, their reliability may suffer from motion artefacts (22). Consequently, validation is required for each foreseen use case. Our group has very recently validated a textile-based electrocardiogram (ECG) and respiratory monitoring system for sleep apnea monitoring (23). The goal of the present study was to assess the reliability and accuracy of a very similar monitoring system for potential integration into a medical early warning system designed to monitor pilots and soldiers. This system was worn by the pilots during low-pressure and high-G exposure training.

## 2. Materials and Methods

In the current study, the accuracy and relevance of employing a textile-based monitoring system for monitoring pilots under extreme conditions were evaluated. The monitoring activities were executed during a low-pressure exposure within a chamber and high-G exposures in a high-performance three-axis centrifuge. For the study, a total of 10 healthy pilot aspirants with an average age of 22.6 years (standard deviation [SD]: 2.2y) volunteered to wear the monitoring system during their regular training and qualification blocks, which were conducted according to standard protocols of the Aeromedical Center. The monitoring of the pilots’ vital parameters (ECG, respiration) were conducted in accordance with the Declaration of Helsinki (24). The Institutional Review Board of the Swiss Aeromedical Center approved the conduct of routine training and qualification modules that are part of pilot education, including the concomitant monitoring of vital signs, and the participants provided written informed consent.

### 2.1. Textile Monitoring System

Throughout the experimental exposures, participants were equipped with a textile monitoring system comprising three distinct sensor types integrated into a belt worn around the chest. The system included two knitted ECG electrodes integrated into the textile assembly for 1-lead ECG recording (along lead I) for the calculation of heart rate (HR) and heart rate variability (HRV) as the SD of RR-intervals (SDNN). A conductive stretch-sensitive textile sensor was applied to measure changes in chest excursion (Figure 1). Based on these measurements, respiratory frequency (fresp) and the respiratory depth (referred to as respiratory sensor elongation amplitude [ampresp]). The third sensor type measured skin head flux in the chest region and was not considered for further analysis in the current study.

**Figure 1:**
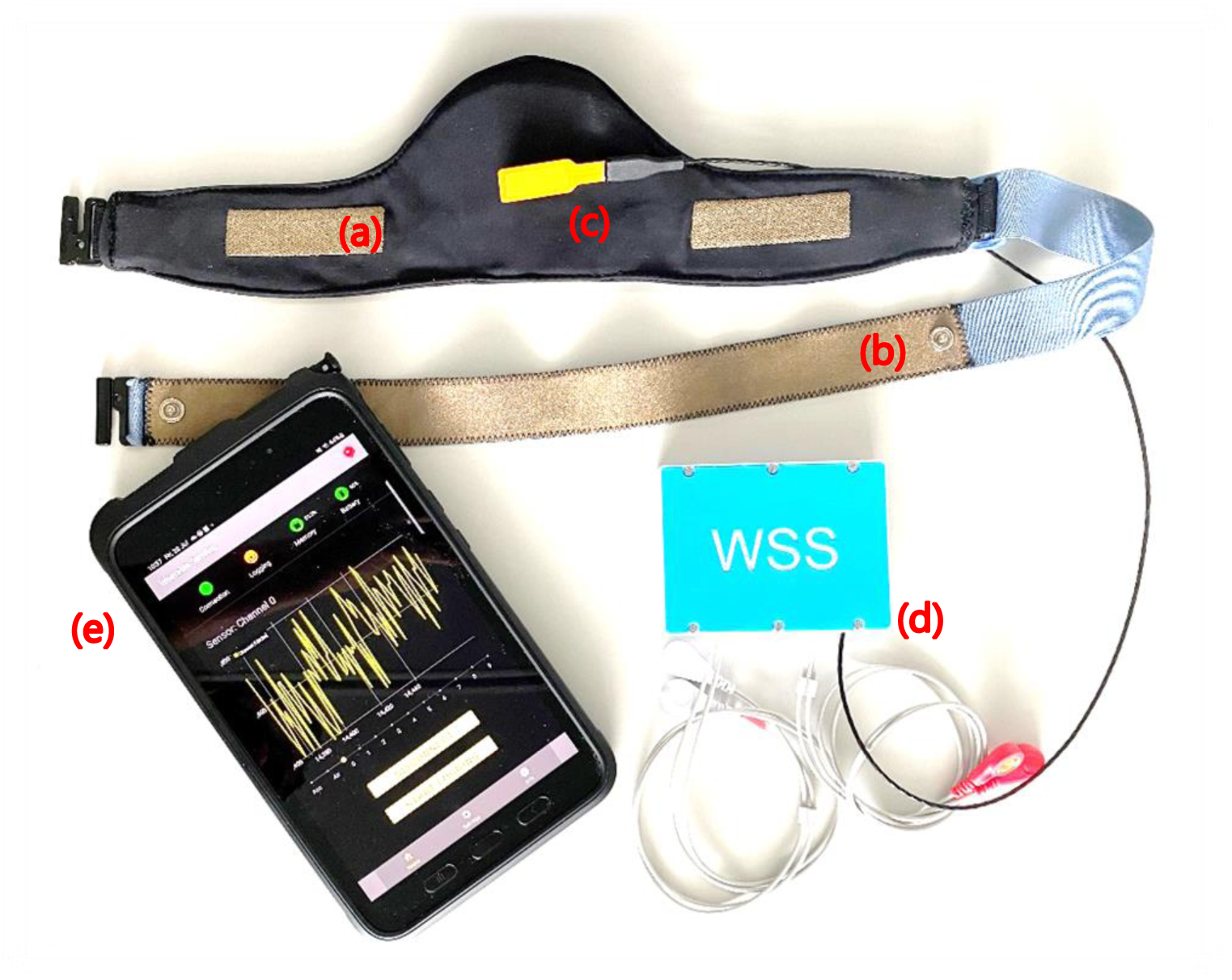
Textile monitoring system consisting of knitted electrodes (a) for ECG signal acquisition, a conductive stretch-sensitive textile (b) for the measurement of chest excursions, and a skin heat flux sensor (c; data not considered for further analysis in this study). The textile monitoring system was connected to a custom-made data logger (d) transferring data to a tablet (e) via Bluetooth.

The 1-lead ECG system was previously clinically validated with 242 patients (Fontana, Martins et al. 2019). A similar combined system of ECG and respiratory monitoring was also used and validated for apnea monitoring and diagnostics (Baty, Cvetkovic et al. 2024). The data was recorded using a custom-made data logger placed in a pocket at the lower abdomen. Respiration was recorded at 25 Hz (Texas Instruments ADS131M08), while the ECG was recorded at 512 Hz (NeuroSky BMD101). Additionally, the logger was equipped with an Inertial Measurement Unit (IMU; Analog Devices ADXL345) to measure the acceleration in all spatial directions. To identify disturbances in signal quality motion artefacts, our group recently developed a method for signal quality assessment that was also applied in this present study (25). The method considers the entire PQRST complex of the ECG signal. The individual QRS and PQRST complexes are averaged for specific time segments of 15s. The similarity between each complex and the average value of the complexes (considered as reference complex for a specific time segment) is calculated using the Pearson correlation analysis. As a third parameter, the standard deviation of the RR-intervals in the measured segments is determined, and the number of outliers is assessed. We defined the plausibility of RR-intervals in a time segment to be true when the standard deviation of RR-intervals was between 1 and 200ms and no more than 2 outliers were detected (i.e. values corresponding to heart rates over 180 or under 30 beats per minute). This allowed a classification of the signal quality into 4 classes, considering the mean values of all Pearson correlation coefficients (PCC) in the ECG time segment and the plausibility criteria (Table 1). Classes 1 and 2 were considered suitable for very accurate analyzes of the RR-intervals.

**Table 1:** Definition of the signal quality classes using the Pearson correlation coefficients (PCC) for the QRS and PQRST complexes of the ECG signal and the plausibility criteria for the RR intervals (25).

| Class | PCC for QRS complexes | PCC for PQRST complexes | Plausibility check |
| --- | --- | --- | --- |
| 1 | $\geq 0.98$ | $\geq 0.98$ | TRUE |
| 2 | $\geq 0.98$ | $< 0.98$ | TRUE |
| 3 | $< 0.98$ | $< 0.98$ | TRUE |
| 4 | any | any | FALSE |

Similarly, the median value of single chest excursion cycles for time segments of 30 seconds duration was calculated to assess the signal quality of respiration. The similarity of chest excursion cycles with the reference median value for a particular time segment was determined based on PCC. The signal was classified into three quality classes based on mean PCC values obtained for respiratory time segments: Class 1: PCC ≥ 0.85, Class 2: 0.75 < PCC < 0.85, and Class 3 ≤ 0.75.

### 2.2. Hypobaric hypoxia exposure

The pilot aspirants underwent a standard training commonly practised at the Aeromedical Institute (FAI Dübendorf, Switzerland) in a low-pressure chamber, simulating an aviation scenario involving a loss of cabin pressure. The subjects participated in groups of two pilots in the exposure sessions, with a medical doctor closely supervising each session. Before the exposure, they underwent preoxygenation using aviation oxygen masks (AOM). Throughout the experiment, the AOM was intermittently removed for specific periods of the protocol (Table 2, Figure 2). During AOM-off periods, the participants were instructed to perform simple calculation tasks and respond to questions from the medical doctor to assess their cognitive performance. The numerical analysis of the data from the wearable system encompassed six distinct data intervals with an identical duration of 30 seconds each, denoted as i1 to i6 (Figure 2). Data interval 1 (i1) captured the timeframe at the end of Phase 2 (ascend with AOM applied, Table 2). Phase 3 consists of the continuing ascent to 7500m asl and staying at this altitude without AOM. Data interval 2 (i2) includes data at the beginning of phase 3, while data interval 3 (i3) consists of data just before the AOM was reapplied (indicating the end of phase 3). Data interval 4 (i4) considers the time after the second removal of the AOM at an altitude of 9000m asl, and data interval 5 (i5) accounts for the 30 seconds just before the AOM was reapplied again at the end of Phase 5. Phases 3 and 5 consisted of hypoxic conditions.

**Figure 2:**
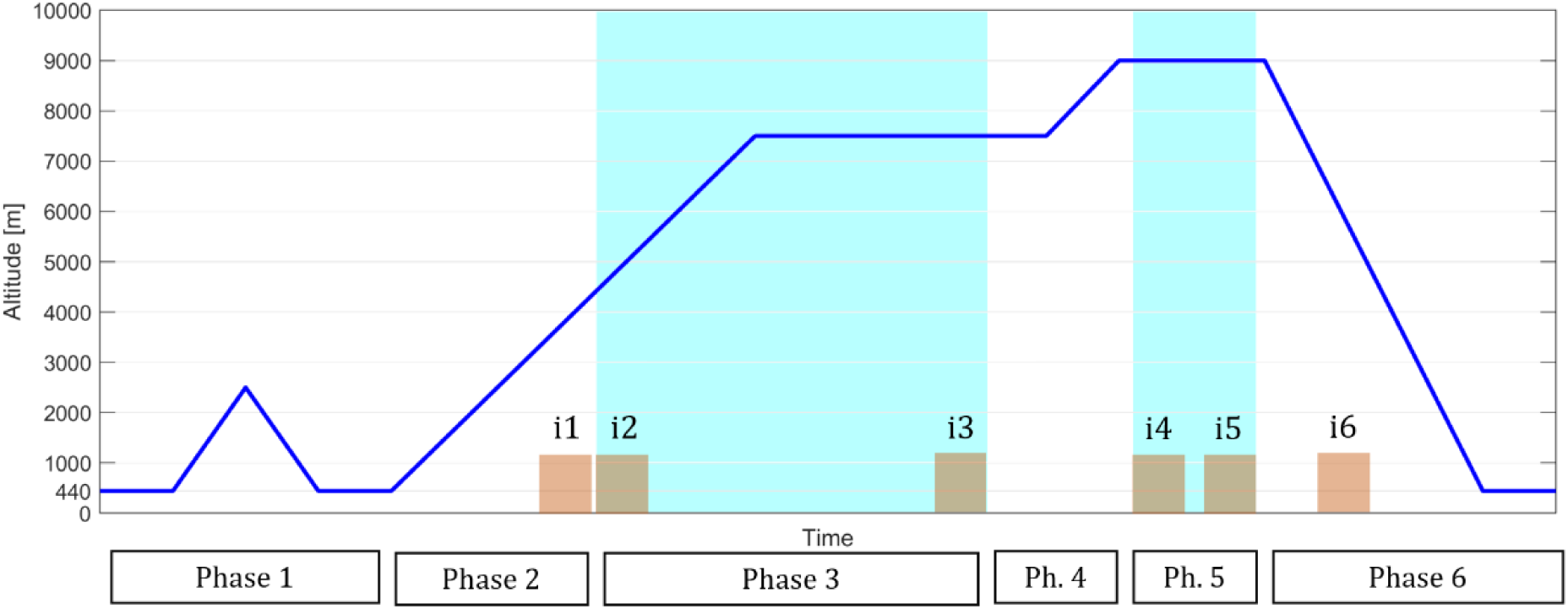
Simulated altitude profile. The blue periods highlight the phases when the participants had their aviator oxygen masks removed (phases 3 and 5). Details regarding the phases are available in Table 2. The orange annotations (i1 to i6) indicate the specific 30-second measurement intervals for ECG and breathing signals.

**Table 2:** Hypobaric hypoxia exposition protocol. The altitude information refers to the altitude that corresponds to the air pressure in the chamber.

|  |  |
| --- | --- |
| Phase 1 | Rise to 2500m above sea level (asl) and back to local altitude (440m asl) with aviation oxygen mask (AOM) on for test of equipment and pressure equalization. |
| Phase 2 | Rise to 4500m asl at 5.55m/s ascent with AOM on. |
| Phase 3 | At an altitude of 4500m asl, the AOM was removed and the participant was instructed to start the calculation exercise. The altitude was gradually increased to 7500m asl, during which the AOM remained off until the medical doctor intervened and put the AOM back on. |
| Phase 4 | Rise to 9000m asl at 5.55m/s ascent with AOM on. |
| Phase 5 | At the altitude of 9000m, the AOM were removed again, and the participants continued the calculation exercise until the medical doctor put the AOM back on. |
| Phase 6 | Decline back to local altitude with AOM on. |

### 2.3. High-G exposures

The participants underwent high-G exposures in a high-performance three-axis centrifuge at the Center for Aerospace Medicine in Königsbrück, Germany, to receive G-force qualification for military jet pilots. For this, pilot aspirants had to successfully complete a 7G exposure for 15s duration (according to NATO STANAG 3827). The 7G exposure was preceded by two exposures of 4G and 5.5G separated by baseline exposures of 1.4G with individual duration between 1 and 2min (Figure 3). The numerical analysis of the data from the wearable system encompassed nine distinct data intervals (i1 – i9) with an identical duration of 15 seconds each, just before, during, and right after high-G exposures (Figure 3).

**Figure 3.**
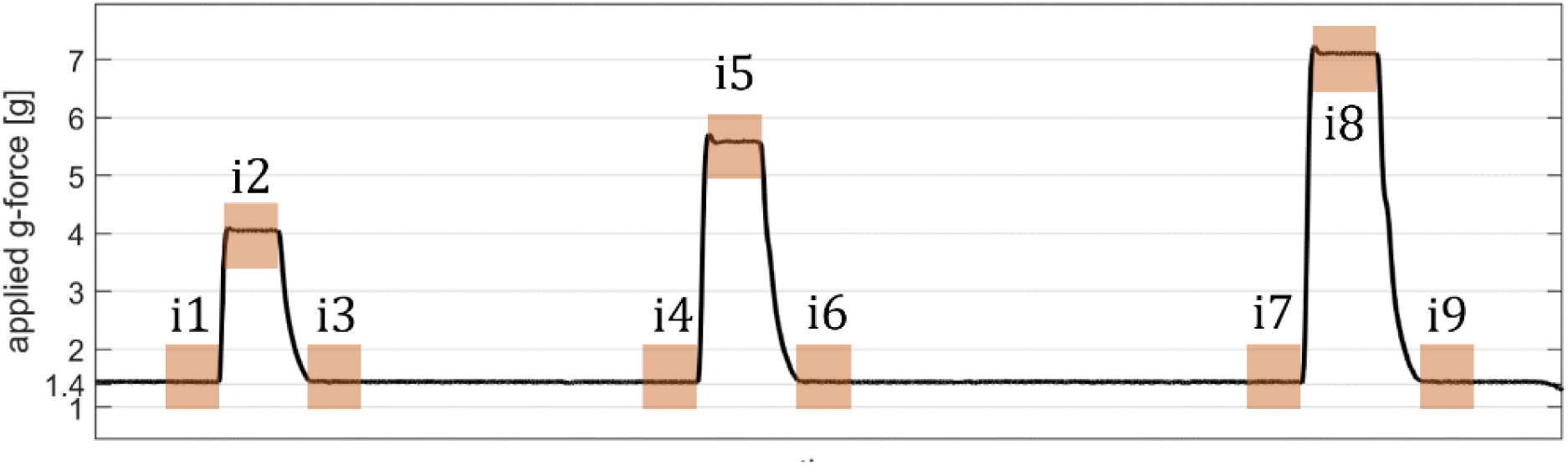
G-force exposure protocol for the qualification profile. The 15-second intervals i1 to i9 were used for the numeric analysis of the wearable system data (ECG and breathing signals). The orange annotations (i1 to i9) indicate the specific 30-second measurement intervals for ECG and breathing signals.

### 2.4. Statistical analysis

A univariate repeated measures ANOVA with post hoc tests corrected according to Tukey was conducted for ECG (HR, HRV) and respiration-related parameters (fresp, ampresp). Data sets were checked for sphericity, homogeneity and normality. Corrections were applied for data sets not complying with sphericity requirements (Greenhouse-Geisser; data sets corrected are indicated in the text). The Jamovi software was used for statistical analysis (26). The level of significance was set at 0.05.

## 3. Results

### 3.1. Hypobaric hypoxia exposures

The quality of the ECG signals obtained during the hypobaric hypoxia exposure of the participants was assessed and classified according to our signal quality assessment approach (Table 1). A total of 91.7% of the ECG time segments were assigned to classes 1 to 3, indicating suitability for the further analysis of HR and HRV. The assessment of respiratory signals identified 71.9% of the respiratory time segments classified as class 1 or 2, which is required for the further analysis of respiratory signals (Table 3).

**Table 3:** The mean ECG (ECG SQ) and respiratory signal quality (Resp SQ) and associated standard deviation (SD) for time segments assigned to classes 1 to 4 (ECG SQ) and classes 1 to 3 (Resp SQ) observed in 10 participants during hypobaric hypoxia exposure (see section 2.1 and Table 1 and for detailed assignment criteria).

| ECG SQ | Mean | SD | Resp SQ | Mean | SD |
| --- | --- | --- | --- | --- | --- |
| Class 1 | 60.2% | 32.2% | Class 1 | 48.5% | 21.5% |
| Class 2 | 22.1% | 17.1% | Class 2 | 23.5% | 9.0% |
| Class 3 | 9.4% | 20.6% | Class 3 | 28.1% | 17.5% |
| Class 4 | 8.3% | 9.2% |  |  |  |

A statistically significant change in HR (repeated ANOVA, Greenhouse-Geisser corrected p<0.001) and HRV (repeated ANOVA, Greenhouse-Geisser corrected p<0.01) was detected (Figure 5). No change in HR (+1%, p=0.98) was observed right after the initial removal of the AOM (i2), while HRV dropped by 22% (p=0.16). At the end of the first hypoxia exposure at 7500m asl (i3), both HR and HRV revealed significant changes by +34% (p<0.001) and -64% (p<0.01), respectively, when compared to i1. The second hypoxia exposure at 9000m asl resulted in a shorter exposure time (i5), while HR (+21%, p=0.07) and HRV (-44%, p=0.157) were not affected significantly when compared to i1, but a significant change was detected when compared to previous data interval i4 (Figure 5). After the application of the AOM (i6), values returned immediately to preexposure data for HR (+2%, p=0.99) and HRV (+25%, p=0.84).

**Figure 5:**
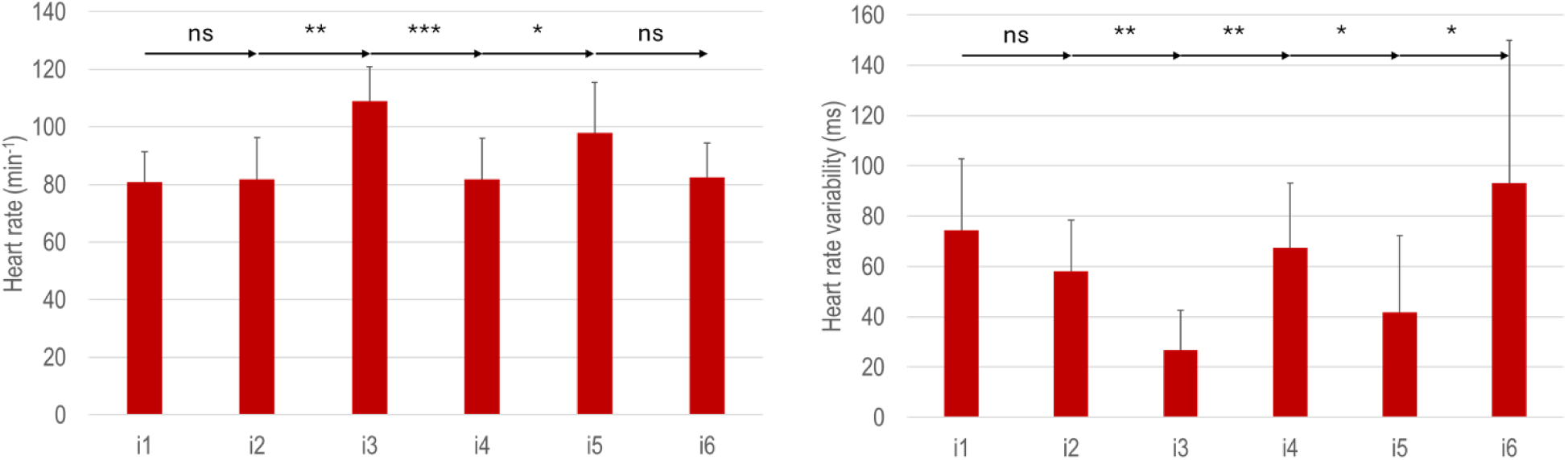
Heart rate and heart rate variability for various measurement intervals (i1 – i6; further information on the prevailing conditions during the individual measurement intervals can be found in Figure 2.) during hypobaric hypoxia exposure (ns: non-significant difference, *: p≤0.05, **: p≤ 0.01, ***: p≤0.001).

Respiratory data did not reveal any statistically significant changes in respiratory frequency (repeated ANOVA p=0.06) nor respiratory sensor elongation amplitude (repeated ANOVA, Greenhouse-Geisser corrected p=0.24; Figure 6). Nevertheless, remarkable changes were observed right after the initial removal of the AOM (i2) for fresp (+18%) and ampresp (-15%) as well as at the end of the first hypoxia exposure at 7500m asl (i3; +26% and -10% for fresp and ampresp, respectively), when compared to i1. The second hypoxia exposure at 9000m asl (i5) revealed similar changes in fresp (+26%), while ampresp was affected more strongly (-20%), when compared to the first hypoxia exposure. After the application of the AOM (i6), values returned immediately to preexposure data for fresp (+0.3%) and showed increased ampresp (+22%).

**Figure 6:**
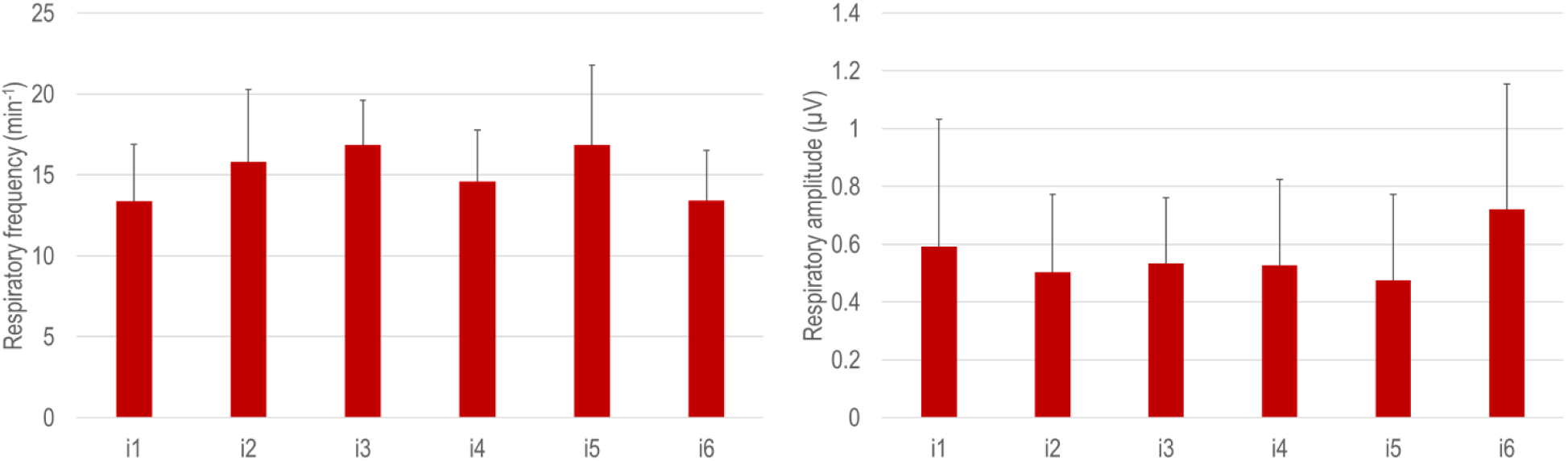
Respiratory frequency and respiratory sensor elongation amplitude for various measurement intervals (i1 – i6; further information on the prevailing conditions during the individual measurement intervals can be found in Figure 2.) during hypobaric hypoxia exposure.

### High G-exposition

Exemplary data is provided for the continuous monitoring of ECG signals and chest excursion (Figure 7). The signal quality was impaired, particularly during the acceleration phases, with most of the signals in class 3 (indicated by the ECG curve colored in orange; Figure 7, left).

**Figure 7:**
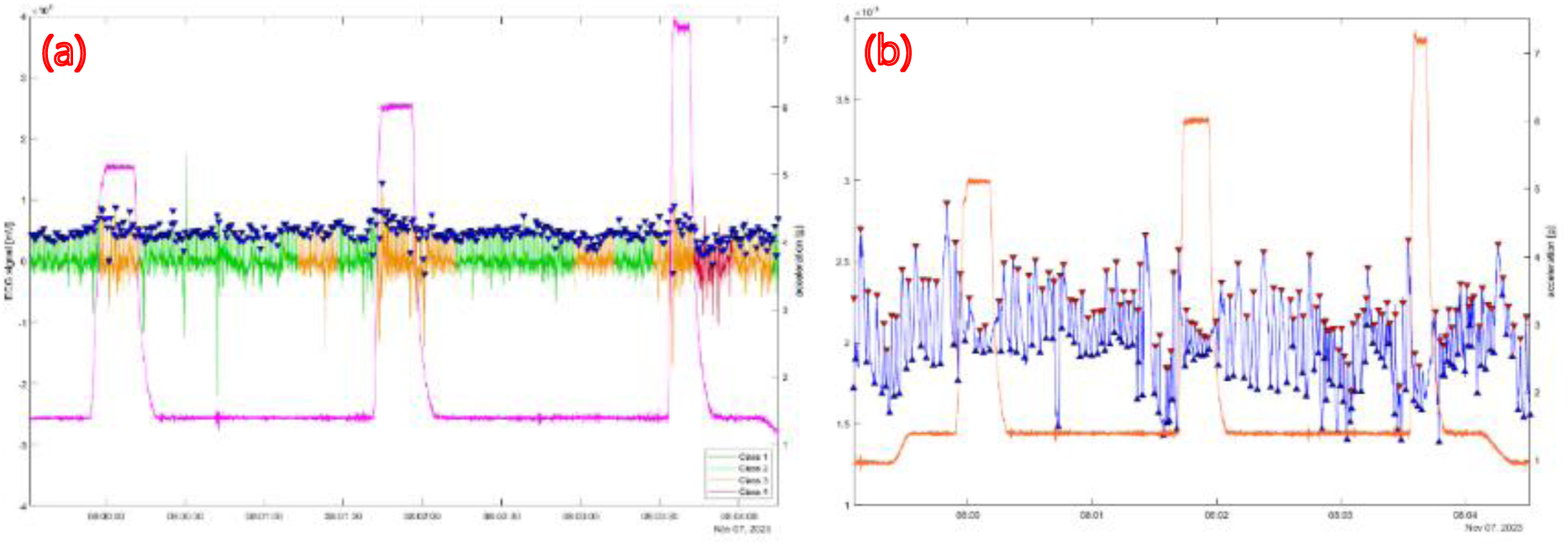
Exemplary data for ECG (a) and chest excursion signals (b) during high G-exposure qualification. The colouration of the ECG signal based on signal quality assigned to time windows of 15s duration: class 1 (dark green), class 2 (light green), class 3 (orange) and class 4 (red). In purple, the acceleration data from the Inertial Measurement Unit is presented. Chest excursion data obtained from respiratory sensor elongation (blue) and acceleration (orange).

A quantified overview on how ECG time segments and respiratory time segments were attributed to the different quality classes is provided in Table 6. A total of 82.8% of the ECG time segments and 76.0% of respiratory time segments were eligible for further analysis.

**Table 6:** The mean ECG (ECG SQ) and respiratory signal quality (Resp SQ) and associated standard deviation (SD) for time segments assigned to classes 1 to 4 (ECG SQ) and classes 1 to 3 (Resp SQ) observed in 10 participants during high-G exposition (see section 2.1 and Table 1 and for detailed assignment criteria).

| ECG SQ | Mean | SD | Resp SQ | Mean | SD |
| --- | --- | --- | --- | --- | --- |
| Class 1 | 1.1% | 3.5% | Class 1 | 47.1% | 18.7% |
| Class 2 | 47.8% | 30.6% | Class 2 | 28.8% | 11.7% |
| Class 3 | 33.3% | 18.1% | Class 3 | 24.0% | 11.6% |
| Class 4 | 17.8% | 19.7% |  |  |  |

A significant change in HR was found during repeated exposures to high G-forces (repeated ANOVA, Greenhouse-Geisser corrected p<0.001), while no effect was observed for HRV (repeated ANOVA p=0.06). During high G-exposures, HR was significantly increased when compared to preceding values (19.5%, 17.5%, and 13.0% for 4G. 5.5G and 7G exposures, respectively (Figure 8). A slight increase was also observed for resting HR at i4 (+4.3%, p=0.41) and i7 (+13.7%, p<0.05). For HRV, a high variation was observed particularly during and right after high G-exposure (Figure 8, right). Particularly during high G-exposure phases, a reduction in ECG signal quality was observed. Mean ECG time segment quality was increased for i2 (2.8±0.4), i5 (3.1±0.6) and i8 (3.5±0.7) when compared to average signal quality class during preceding data intervals (2.1 – 2.5).

**Figure 8:**
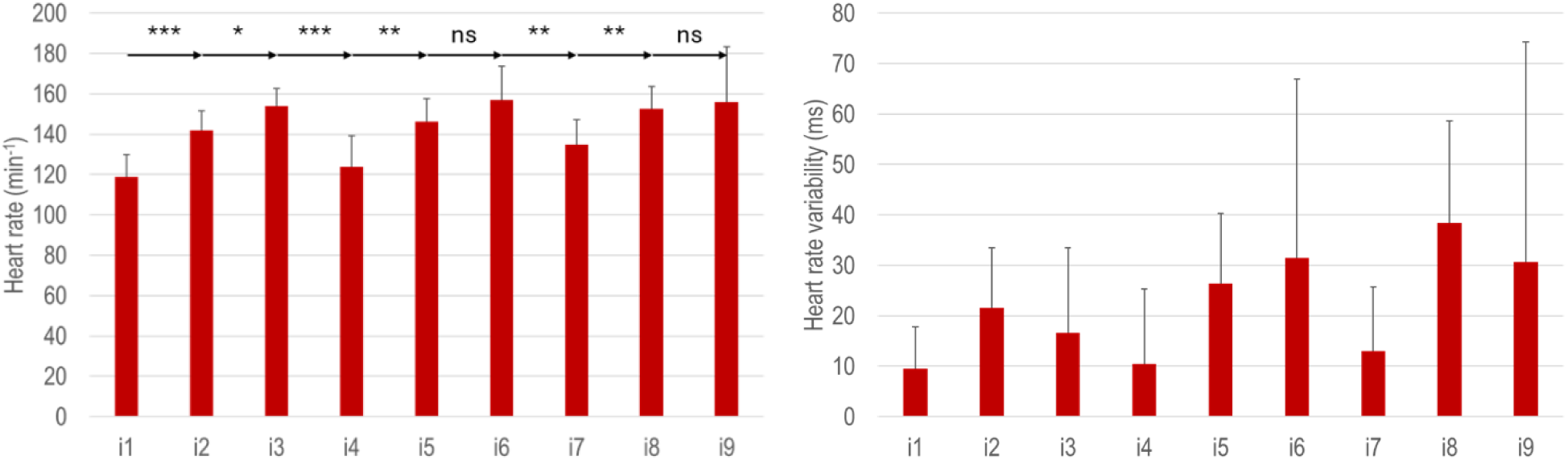
Heart rate and heart rate variability for various measurement intervals (i1 – i9; further information on the prevailing conditions during the individual measurement intervals can be found in section 2.3, Figure 3) during high G-exposures (ns: non-significant difference, *: p≤0.05, **: p≤ 0.01, ***: p≤0.001).

Even though a statistically significant effect was detected for fresp (repeated ANOVA p=0.007), no significant differences between subsequent data intervals were identified. A high coefficient of variation was particularly detected during measurement intervals right after high G-exposures (26.1%, 25.5%, and 22.6%, after 4G-, 5.5G-, and 7G-exposure, respectively), which might be attributed to a high average signal quality class for respiratory signals during respective measurement intervals (2.1±0.9, 1.7±0.7, and 1.9±0.7 for 4G-, 5.5G-, and 7G-exposure, respectively). In contrast, fresp variation was found to be much lower during high G-exposure measurement intervals with forced respiration (14.2%, 9.5%, and 3.8% during 4G-, 5.5G-, and 7G-exposure, respectively). Also, during these intervals, average signal quality classes were found to be improved when compared to above values (1.9±0.9, 1.3±0.8, and 1.9±1.1 for 4G-, 5.5G-, and 7G-exposure, respectively).

## 4. Discussion

The textile monitoring system investigated achieved 91.7% and 82.2% of ECG time segments meeting the plausibility criteria (signal quality classes 1-3) during hypobaric hypoxia and high G-force exposure, respectively. For respiration, 71.9% and 76.0% of plausible respiratory time segments were identified during hypobaric hypoxia and high G-force exposure, respectively. In the low-pressure chamber, 82.3% of ECG time segments were classified as class 1 or 2, while this value was found to be 48.9% in the high-performance centrifuge only. Particularly during high G-exposures, heart rate variability was found to be increased even though a reduction in this variable would have been expected due to increased physical stress. We, therefore, assume that the ECG signal was affected by an increased muscular tension (mainly legs and torso), and the Valsalva manoeuvres resulted in increased muscular activity in the torso. This affected the electrophysiological recording of the ECG signal (i.e. superimposition of the ECG signal with increased electromyographic activity). Furthermore, the exposure to high accelerations in the centrifuge might also lead to motion artefacts. As a result, a reduction in signal quality was particularly observed during high G-exposure phases. In contrast, no increased muscular activity of legs and torso is expected during hypobaric hypoxia exposure (not measured in this study). Pilot aspirants were sitting in the low-pressure chamber without any accelerations. The calm body posture induced less noise and resulted in a higher data quality. Consequently, only 8% of the ECG time segments were classified in class 4 during hypoxia exposure and had to be discarded for further analysis.

Despite the additional muscular activity and additional movement and force-related implications, only 18% of the ECG time segments monitored with our textile monitoring system had to be discarded for further analysis.

The respiratory signals recorded with the same textile monitoring system did not reveal significant differences between the two training modalities for class 1 (48.5% ± 21.5% during hypoxia exposure and 47.1% ± 18.7% during high G-exposure) and class 2 (23.5% ± 9.0% and 28.8% ± 11.7% for hypoxia and high-G exposure, respectively). This was somewhat surprising as slightly reduced signal qualities were expected during high-G exposures due to movement artefacts and changes in respiratory patterns, particularly a reduction in respiratory amplitude (Figure 9). This proves that the respiratory sensor measured chest excursions reliably, irrespective of increased muscular activity in the torso or motion artefacts.

**Figure 9:**
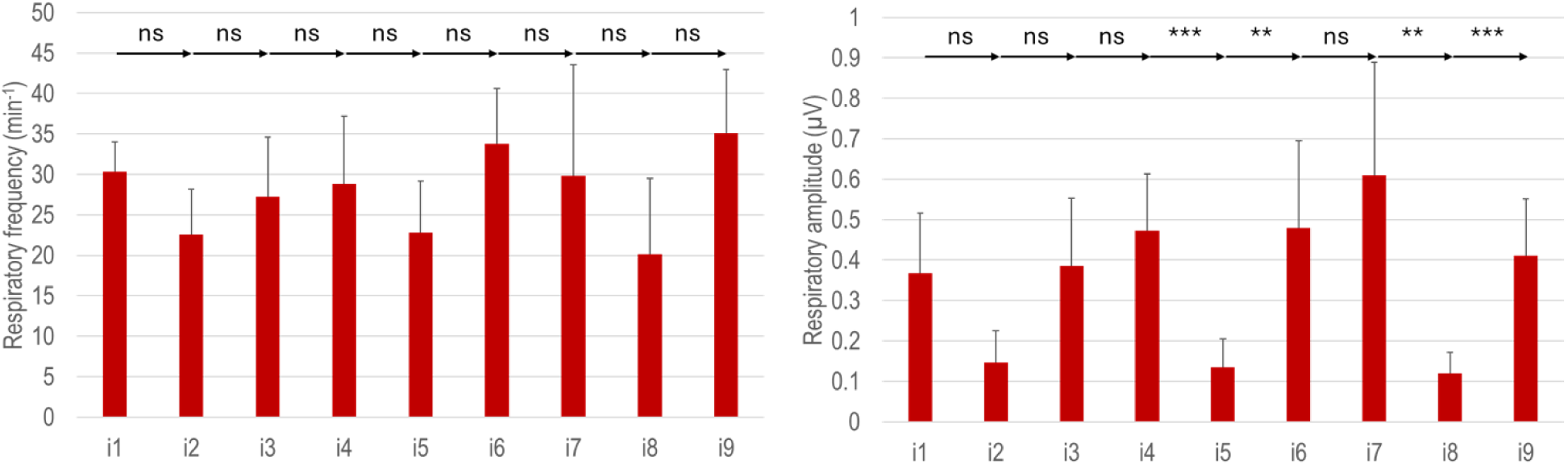
Respiratory frequency and respiratory sensor elongation amplitude for various measurement intervals (i1 – i9; further information on the prevailing conditions during the individual measurement intervals can be found in Figure 3) during high G-exposures (ns: non-significant difference, **: p≤ 0.01, ***: p≤0.001).

Nevertheless, a high inter-participant variability was observed for the accuracy of data acquisition, resulting in standard deviations reaching 20%-30% for the ECG signal quality in both training modalities and around 10%- 20% for the respiratory signal quality. Thus, we assume individual factors, such as the fitting of the monitoring system and skin properties, affect the measurement quality and need to be considered as well.

During the two high-altitude phases simulating 7500 and 9000 m without AOM, the heart rates increased significantly, accompanied by a significant drop in HRV, which is in line with findings from previous studies (6). For the respiratory patterns, the highest respiratory frequencies were found at the end of the hypoxia phases (i3 and i5); however, these changes did not reach statistical significance. Respiration immediately normalised after the application of the AOM, indicated by a reduction in respiratory frequency and an increase in respiratory amplitude. At the same time, an increase in HRV was also measured during interval 6, which shows that pilot aspirants were able to recover very quickly from the hypoxic stress with the administration of oxygen. As heart rate variability was immediately affected by hypoxic exposure and also heart rate revealed significant changes, it may be concluded that these parameters can be used to monitor the physiological stress induced by hypoxia, while respiratory patterns are not clear indicators.

During the high-G exposures, significant differences in heart rate and respiratory amplitude were found. The changes in respiratory patterns were expected due to the breathing manoeuvres to maintain cerebral blood flow, which significantly reduced both the respiratory amplitude and slightly affected respiratory frequency as well. The increase in HR could be expected as well, as this was already demonstrated in several studies (e.g (27). The increase of HR was not related to the level of acceleration, as the mean value reached 154±9 bpm, 157±17 bpm and 152±11 bpm after 4G, 5.5G and 7G exposure, respectively. Peak HR was proposed as a possible predictor of G tolerance during training, as it was dependent on the age, the flying experience and the training status of the pilot aspirants (28,29). The increase in HRV might be observed due to the reduction in signal quality, as discussed above, or due to cardiac arrhythmias, which might have occurred during high-G exposure periods. Nevertheless, the results were found the be in the range reported in a recent study, which concluded that the significant change in HRV during a 5.5G exposure shows that this parameter may be used as a stress predictor and indicator for G-tolerance (30).

## 5. Conclusion

The main goal of this study was to analyse the feasibility of using a textile-based sensor system for the monitoring of military pilots during different training modalities, such as hypobaric hypoxia, high-G exposure training or qualifications. The sensors were integrated into a textile belt that was worn around the chest. The ECG signals could be measured with a very good signal quality: over 90% of the ECG time segments allowed for further analysis of the signals during hypoxia training and over 80% during high-G-force training, which makes them suitable for very accurate analyses of the RR intervals. The recording of combined physiological data, such as ECG and respiratory signals, allows a more holistic interpretation of body responses when exposed to extreme and even hazardous environmental conditions. The effect of hypoxia could clearly be shown by immediate changes in heart rate variability and following changes in heart rate, while the respiratory patterns were less conclusive. During high G-force exposure, respiratory patterns revealed, such as a decrease in frequency and amplitude, in combination with changes in heart rate, provide objective information about the physical stress the pilot aspirant was exposed to. Peak heart rate as well as HRV can be used as indicators of the G-tolerance of the pilot aspirants.

The applicability of such textile-based sensors for continuous monitoring of vital signs of military pilots during training was demonstrated. Further studies are planned with the inclusion of additional vital signs like oxygen saturation and body temperature. Such sensors can be potentially directly integrated into the pilot’s clothing. The collected data will be complemented with data from further studies and used to establish an algorithm for the early prediction of possible physical and cognitive impairments of pilots during training and on-duty flights.

## Data Availability

Data cannot be shared publicly because the written consent for further use of data in encrypted form is not available for all the study participants. Data are available from the Institutional Data Access (contact via) for researchers who meet the criteria for access to confidential data.

## Acknowledgements

We sincerely thank all pilot aspirants for their valuable contribution to this study.

